# Evolution of immunity to SARS-CoV-2

**DOI:** 10.1101/2020.09.09.20191205

**Authors:** Adam K. Wheatley, Jennifer A. Juno, Jing J. Wang, Kevin J. Selva, Arnold Reynaldi, Hyon-Xhi Tan, Wen Shi Lee, Kathleen M. Wragg, Hannah G. Kelly, Robyn Esterbauer, Samantha K. Davis, Helen E. Kent, Francesca L. Mordant, Timothy E. Schlub, David L. Gordon, David S. Khoury, Kanta Subbarao, Deborah Cromer, Tom P. Gordon, Amy W. Chung, Miles P. Davenport, Stephen J. Kent

**Affiliations:** Department of Microbiology and Immunology, University of Melbourne, at The Peter Doherty Institute for Infection and Immunity, Melbourne, Victoria, Australia; Australian Research Council Centre for Excellence in Convergent Bio-Nano Science and Technology, University of Melbourne, Melbourne, Victoria, Australia; Department of Immunology and Flinders Proteomics Facility, College of Medicine and Public Health, Flinders University, Adelaide, South Australia, Australia; Kirby Institute, University of New South Wales, Kensington, New South Wales, Australia; Melbourne Sexual Health Centre and Department of Infectious Diseases, Alfred Hospital and Central Clinical School, Monash University, Melbourne, Victoria, Australia; Sydney School of Public Health, Faculty of Medicine and Health, University of Sydney, Sydney, New South Wales, Australia; Department of Microbiology and Infectious Diseases, Flinders University and SA Pathology, Flinders Medical Centre, Adelaide, South Australia, Australia; WHO Collaborating Centre for Reference and Research on Influenza, The Peter Doherty Institute for Infection and Immunity, Melbourne, Victoria, Australia; Department of Immunology, SA Pathology, Flinders Medical Centre, Adelaide, South Australia, Australia

**Author notes:** Corresponding author. Address to Professor Stephen Kent, Department of Microbiology and Immunology, Doherty Institute, 792 Elizabeth St, Melbourne, Victoria, Australia 3000. AKW, JAJ and JJW contributed equally.

## Abstract

The durability of infection-induced SARS-CoV-2 immunity has major implications for public health mitigation and vaccine development. Animal studies^1,2^ and the scarcity of confirmed re-infection^3^ suggests immune protection is likely, although the durability of this protection is debated. Lasting immunity following acute viral infection requires maintenance of both serum antibody and antigen-specific memory B and T lymphocytes and is notoriously pathogen specific, ranging from life-long for smallpox or measles^4^, to highly transient for common cold coronaviruses (CCC)^5^. Neutralising antibody responses are a likely correlate of protective immunity and exclusively recognise the viral spike (S) protein, predominantly targeting the receptor binding domain (RBD) within the S1 sub-domain^6^. Multiple reports describe waning of S-specific antibodies in the first 2-3 months following infection^7-12^. However, extrapolation of early linear trends in decay might be overly pessimistic, with several groups reporting that serum neutralisation is stable over time in a proportion of convalescent subjects^8,12-17^. While SARS-CoV-2 specific B and T cell responses are readily induced by infection^6,13,18-24^, the longitudinal dynamics of these key memory populations remains poorly resolved. Here we comprehensively profiled antibody, B and T cell dynamics over time in a cohort recovered from mild-moderate COVID-19. We find that binding and neutralising antibody responses, together with individual serum clonotypes, decay over the first 4 months post-infection, as expected, with a similar decline in S-specific CD4+ and circulating T follicular helper (cTFH) frequencies. In contrast, S-specific IgG+ memory B cells (MBC) consistently accumulate over time, eventually comprising a significant fraction of circulating MBC. Modelling of the concomitant immune kinetics predicts maintenance of serological neutralising activity above a titre of 1:40 in 50% of convalescent subjects to 74 days, with probable additive protection from B and T cells. Overall, our study suggests SARS-CoV-2 immunity after infection is likely to be transiently protective at a population level. SARS-CoV-2 vaccines may require greater immunogenicity and durability than natural infection to drive long-term protection.

---

We recruited a longitudinal cohort of 64 subjects who recovered from COVID-19 (Extended Data Fig 1). A total of 158 samples were collected between day 26 and 149 post-symptom onset, with samples nominally denoted as early (≤50days), intermediate (50-100 days) and late (≥100 days) convalescence (Fig 1A). In early convalescence, neutralisation activity was widespread with a median serological titre of 52, which declined to 34 in late convalescence (Fig 1B). A mixed-effects modelling approach found that a two-phase decay model best fit with the observed decay of neutralisation titres across the cohort (p < 0.00001, likelihood ratio test), with rapid decay evident over the first half of our time-series (half-life (t_1/2_) prior to day 70 = 55 days), compared with slower decay in the second half (t_1/2_ from day 70 = 519 days)(Fig 1B). The capacity of immune plasma to inhibit interaction of the SARS-CoV-2 receptor binding domain (RBD) with soluble hACE2 receptor^19^ waned with a similar two-phase decay, dropping more rapidly before day 70 (t_1/2_ = 238 days) and slowing after day 70 (t_1/2_ = 1912 days; Fig1C).

**Figure 1.**
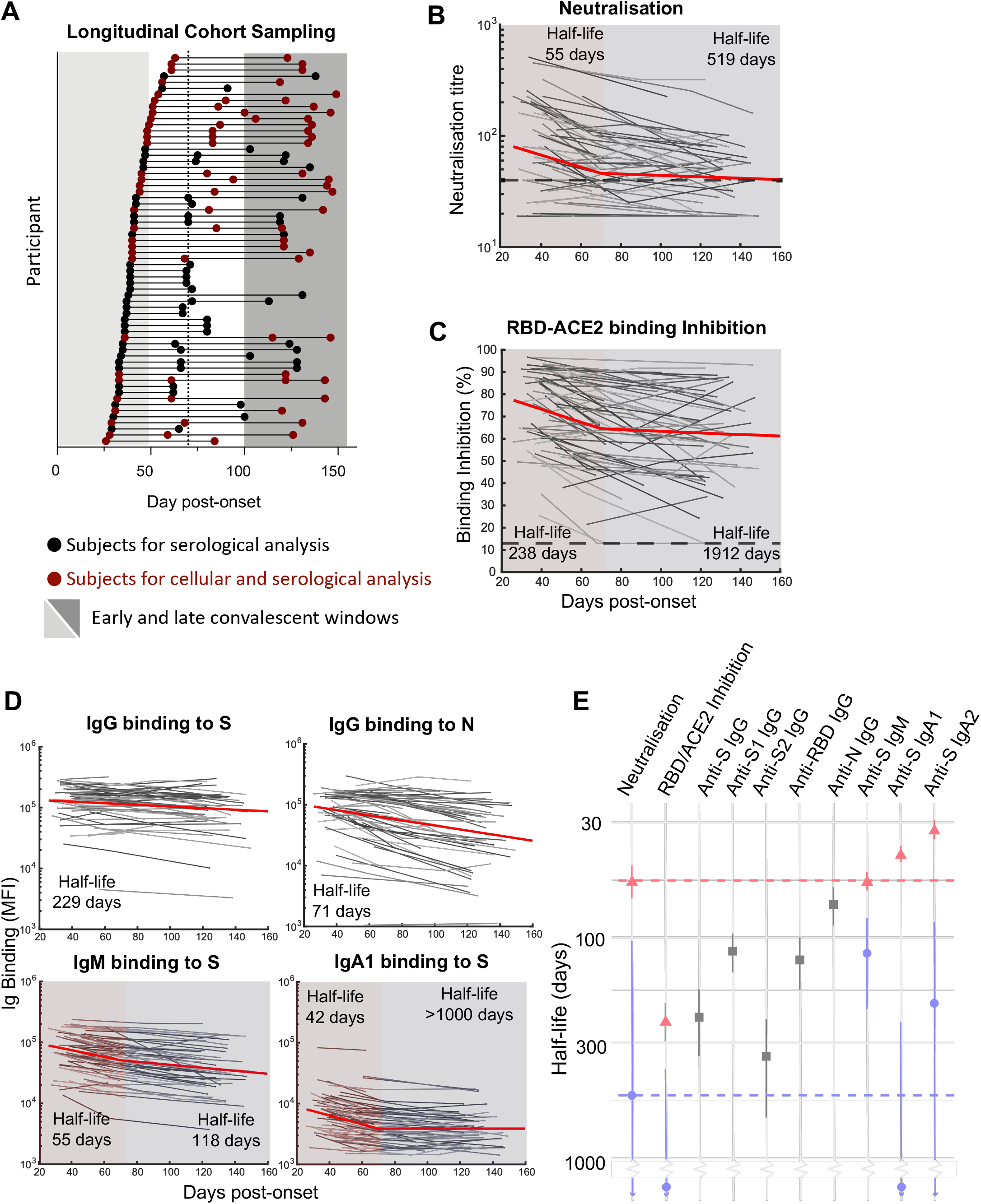
Dynamics of serological responses to SARS-CoV-2.

(**A**) Timeline of sample collection for each cohort participant (n = 64 participants, 158 total samples). Samples included only in serological analysis are indicated in black (n = 33); samples included in both serological and cellular immune analysis are indicated in red (n = 31). Shaded areas indicate early (< 50 days) and late (>100 days) convalescent time periods, and dashed line indicates day 70 midpoint. **(B)** Longitudinal microneutralisation endpoint titre and **(C)** inhibition of ACE2 binding (%) for individuals. Best fit two-phase decay slope (red line) is indicated. **(D)** Individual kinetics and best fit decay slopes for IgG binding to spike (S), IgG binding to nucleoprotein (N), IgM binding to S and IgA1 binding to S. **(E)** Estimated half-life and confidence intervals of the neutralising antibody titre before day70 (red) and after day70 post-symptom onset (blue) are indicated as dashed vertical lines. Estimated early decay rates and confidence intervals for serological inhibition of ACE2 and antibody binding titres are indicated (single phase decay is shown in grey, two phase decay indicated in red/blue).

Plasma antibodies specific for SARS-CoV-2 S antigens (trimeric spike protein (S), S1, S2 and RBD subdomains) and nucleocapsid (N) antigens were quantified longitudinally using a multiplex bead array^25^. In contrast to neutralisation titres, decay of S-specific IgG was best fit by a model of constant decay over the period of observation (t_1/2_ = 229 days), with rates of decay divergent for antibodies binding S1 (t_1/2_ = 115 days), S2 (t_1/2_ = 344 days) and RBD antigens (t_1/2_ = 126; Fig 1D, Extended Data Fig. 2). Kinetics of decay were broadly consistent between IgG1 and IgG2 subclasses, with IgG3 displaying a more rapid, two phase decline (Extended Data Fig. 2). Consistent with a previous report^26^, we find N-specific IgG decays significantly more rapidly than S-specific IgG (t_1/2_ = 71 and 229 days respectively, p < 0.00001, Fig 1D). In contrast to IgG, S-specific IgM and IgA1 fit a two-phase decay, with a more rapid early decay (t_1/2_ = 55 and 42 days respectively) followed by a slower decay in late convalescence (t_1/2_ = 118 and > 1000 days respectively; Fig 1D). A comparison of decay rates between neutralising activity and antibody binding demonstrated that early neutralisation decay occurs at a similar rate to the early decline in S, RBD and S1-specific IgM (Fig 1E, Extended Data Fig 2). Neutralisation titre at both early and late convalescence was well correlated with serum inhibition of RBD-ACE2 binding and S, S1 and RBD specific IgG, IgM (and to a lesser extent IgA1) responses, as well as with S2 and N specific IgG responses (Extended Data Fig 3). Neutralising activity during early convalescence was the best correlate of long-term maintenance of neutralisation responses (Spearman rho = 0.88, p< 0.00001; Extended Data Fig 3). Serum inhibition of RBD-ACE2 binding and S1-specific IgG responses in early infection were also well correlated with neutralisation titre in late convalescence (Spearman rho = 0.79, 0.81, respectively; Extended Data Fig 3). However, in a multiple regression model, once early neutralisation activity was included no other significant predictors were identified (p>0.15 for all other variables).

**Figure 2.**
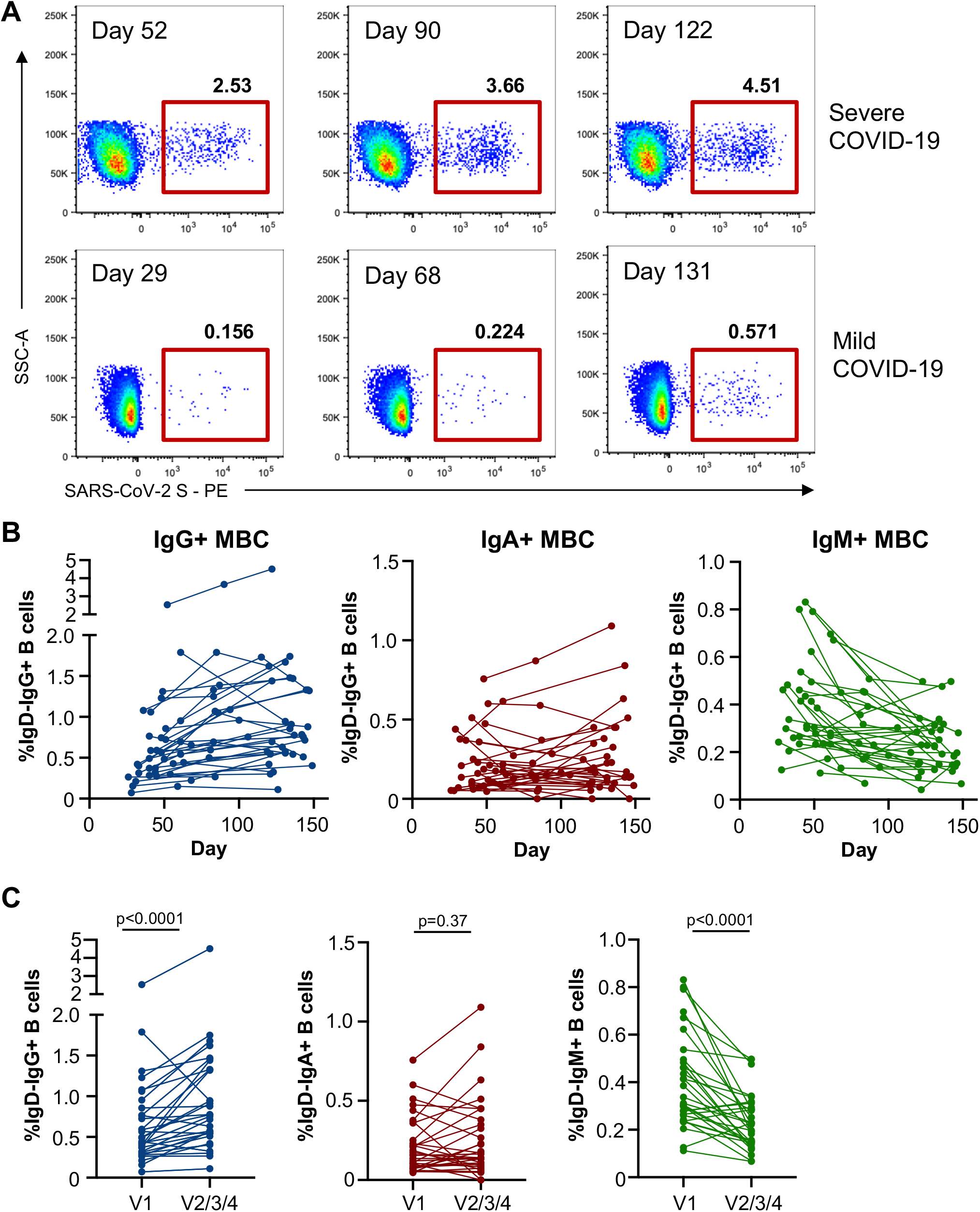
Quantification of S-specific memory B cell responses.

(**A**) Staining class-switched B cells (CD19+IgD-) with SARS-CoV-2 spike probes allows the tracking of antigen-specific cells in subjects previously infected with SARS-CoV-2. (**B**) Frequencies of S-specific IgG+, IgA+ or IgM+ memory B cells as a proportion of CD19+CD20+IgD- B cells in PBMC samples were assessed longitudinally (n = 31 subjects). (**C**) Comparison of S-specific IgG+, IgA+ or IgM+ memory B cell frequencies at the earliest and latest timepoint available for each individual (n = 31). Statistics assessed by two-tailed Wilcoxon test.

**Figure 3.**
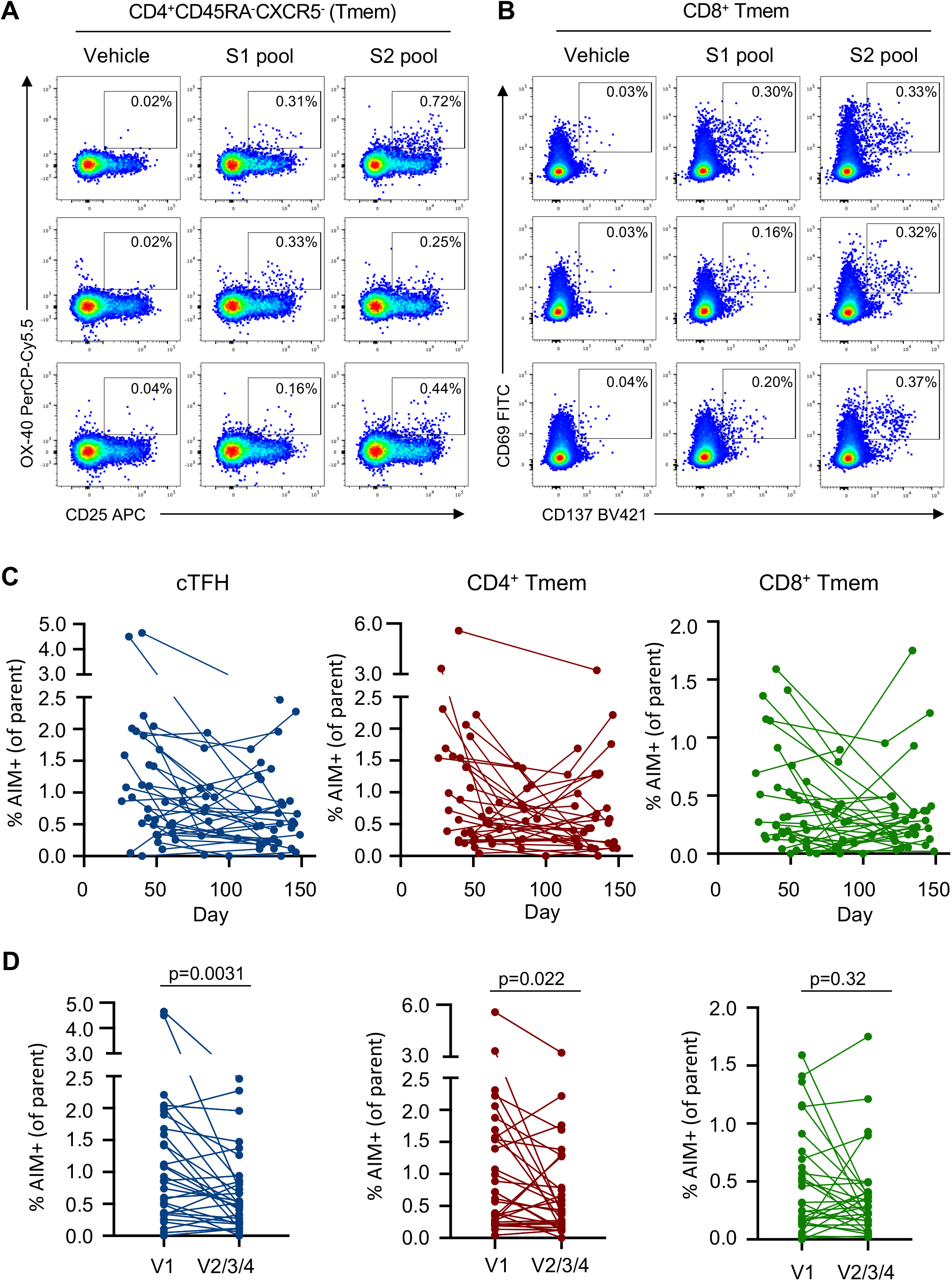
Quantification of antigen-specific CD4+ and CD8+ T cell responses.

(**A**) Representative staining of AIM markers (CD25, OX-40) on CD4+ Tmem cells (CD3+CD4+CD8-CD45RA-CXCR5-) after stimulation with vehicle, S1 or S2 peptide pools in longitudinal samples from 1 participant (top row, day 33; middle row, day 61; bottom row, day 143). (**B**) Representative staining of AIM markers (CD69, CD137) on CD8+ Tmem cells (CD3+CD8+CD4-non-naïve) in longitudinal samples from 1 participant (top row, day 41; middle row, day 85; bottom row, day 120). (**C**) Longitudinal changes in the frequency of total S (S1+S2 pool responses after background subtraction)-specific responses among cTFH, CD4+ and CD8+ Tmem subsets (n = 31). (**D**) Comparison of S-specific T cell responses at the earliest and latest timepoint available for each individual (n = 31). Statistics assessed by two-tailed Wilcoxon test.

The decay of polyclonal antibody in plasma may obscure a more complex picture of the dynamics of individual antibody specificities. To resolve longitudinal serological decay at the level of a single clonotype, we adapted a novel mass spectrometry (MS)-based quantitative proteomics workflow developed for serum autoantibody profiling^27,28^ to track unique CDR-H3 peptides matching recovered S-specific immunoglobulins sequences from convalescent subjects^19^ (n = 4; Extended Data Fig 4A, B). Consistent with the decay of polyclonal S-specific antibody in the blood, we find a decline in the relative abundance over time for each unique clonotype (Extended Data Fig. 4C), although absolute rates of decay did vary, suggesting the kinetics might to some degree be clonotype-, epitope- or subject-specific.

**Figure 4.**
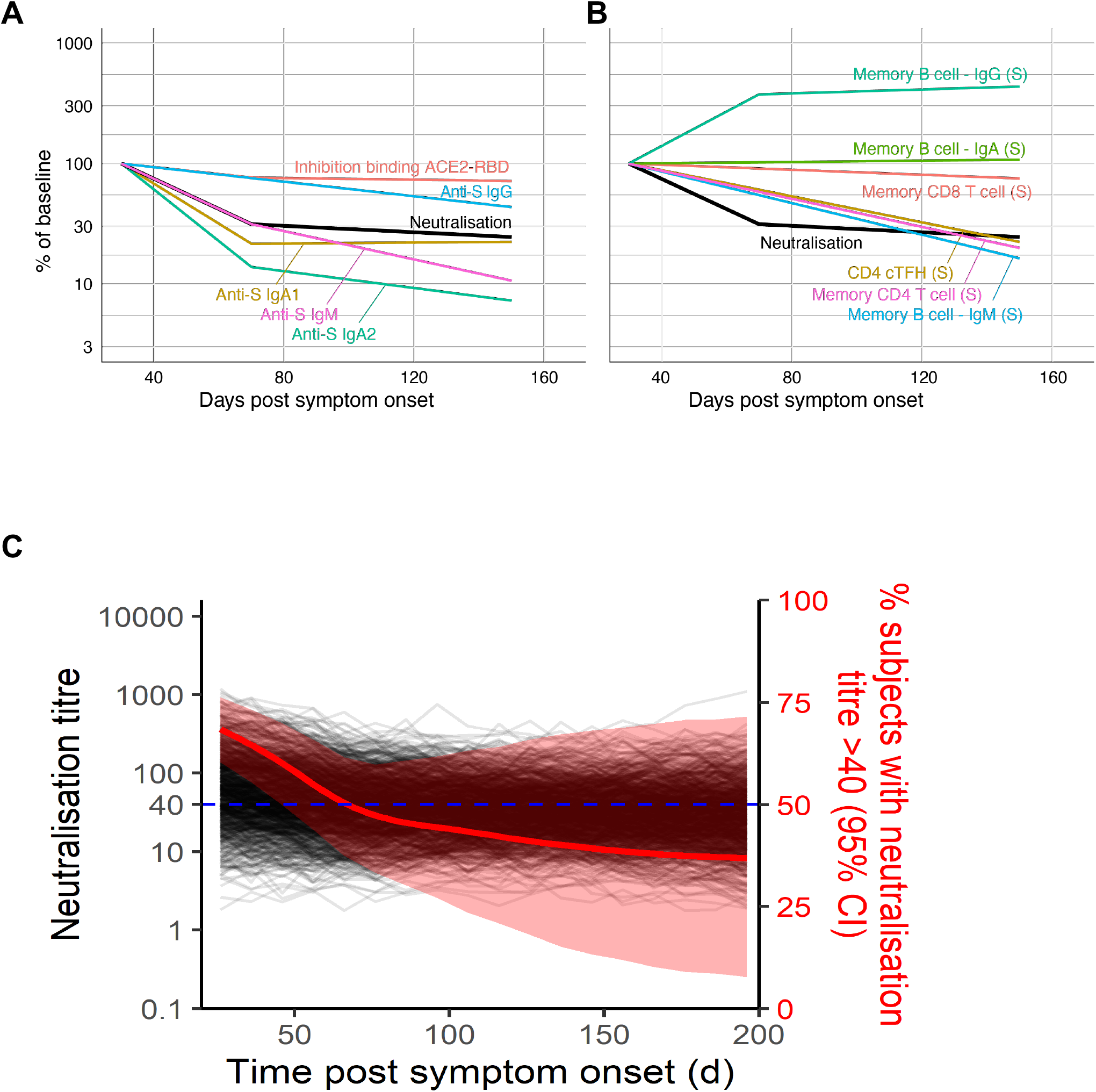
Modelling of concomitant immune responses after COVID-19.

(**A**) Rates of decay of serological neutralisation activity, ACE2 binding inhibition, and S-specific IgG, IgM and IgA following recovery from SAR-CoV-2 infection. (**B**) Fitted Growth and decay rates for S-specific memory T cell and B cell frequencies in PBMC. (**C**) Simulation of elicitation and decay of serological neutralisation activity in 1000 individuals based on distributions observed in our SARS-CoV-2 convalescent cohort. The simulation was repeated 1000 times to estimate the proportion of individuals maintaining a neutralisation titre above 1:40 across multiple simulations (median and 95% confidence intervals shown in red).

Anti-viral memory B and T cell responses will likely make additive contributions to long-term immunological protection against COVID-19. SARS-CoV-2-specific B cell responses were measured longitudinally in 31 subjects where sufficient cells were available (Fig 1A) using flow cytometry and fluorescent S and RBD probes as previously described^19^. Following infection, frequencies of IgG+ S-specific memory B cells increased over time irrespective of disease severity (Fig. 2A and 2B; gating in Supplementary Fig. S1). In contrast, S-specific IgA+ MBC frequencies remained relatively stable while IgM+ MBC frequencies decreased (Fig. 2B). IgG+ S-specific MBC remain significantly elevated at the final relative to the first available sampling (p< 0.0001), contrasting with stable IgA+ (p = 0.367) and declining IgM+ populations (p< 0.0001) (Fig. 2C). Assessment of the activation status of S-specific IgG+ MBC using CD21/CD27 staining^29^ demonstrated decreased proportions of “activated” MBC and a return to a resting (CD27+CD21+) phenotype over time (Extended Data Fig 5). Although present at comparatively low frequencies, the dynamics of RBD-specific MBC largely mirrored that of the parental S-specific population (Extended Data Fig 6). Modelling the growth rates reveals IgG+ S-specific MBC frequencies had a doubling time of 48 days in early convalescence, after which point the doubling time slowed to 843 days, with IgG+ RBD-specific cells MBC broadly comparable (doubling time = 58 days early, t_1/2_ = 247 days late) (Extended Data Fig 7). The consistent and sustained increase in S-specific IgG+ MBC frequencies over time aligns with a prior report of SARS-CoV-2 convalescent subjects^13^, and with reports from other viral infections^30^ or replicating viral vaccines^31,32,6^. Given the relatively low level of somatic mutation observed in S-specific antibodies recovered from convalescent subjects to date^6,19^ comparative studies at 6-9 months post-infection will be informative to understand the maturation of the humoral response over time and the protective potential of stably retained MBC populations.

Anti-viral memory T cell responses have been associated with amelioration of disease for respiratory infection such as influenza^33^. S-specific cTFH and conventional CD4+ and CD8+ memory T cells (Tmem), were quantified using activation induced marker (AIM) assays^19,22^ (Methods) following stimulation with overlapping S (split into S1 or S2) peptide pools (Fig. 3A, 3B). Frequencies of S-specific memory T cells were dynamic over time and varied between individuals, with evidence of either rapid decline or stable maintenance (Fig. 3C). Pairwise comparison of cTFH or CD4+ Tmem frequencies at the final visit relative to the first available sampling demonstrated a significant reduction in S-specific responses over time (p = 0.0031 for cTFH, p = 0.0224 for CD4+ Tmem; Fig. 3D). In contrast, frequencies of S-specific CD8+ Tmem were stable at a population level (p = 0.3247), although individual responses were varied (Fig. 3D). Modelling of the decay rates estimated t_1/2_ of 128 days for cTFH (95% CI 67, 1247) and 119 days for CD4+ Tmem (95% CI 66, 612; Extended Data Fig 8). In contrast, the estimated decay of CD8+ responses is not significantly different from 0 (t_1/2_ = 670 days, 95% CI 97, –136; Extended Data Fig 8).

Therefore, while we and others^34,35^ find that CD4+ responses are generally higher during early convalescence, CD8+ T cell responses appear relatively stable during late convalescence.

Multiple studies have reported CCC cross-reactive CD4+ T cells in SARS-CoV-2 uninfected subjects, largely recognising epitopes in S2^22,35,36^. To understand the influence that recall of pre-existing CCC cross-reactive immunity might have on decay, we contrasted S1 and S2 responses among the CD4+ T cell subsets. For cTFH, a significant drop in S1 responses was observed over time (p = 0.0028), while S2 responses were comparably stable but did similarly trend downward (p = 0.0657) (Extended Data Fig 9A,B). Analogous patterns were observed for the CD4+ Tmem cells (Extended Data Fig 9C,D). Consequently, S2-specific cTFH and CD4+ Tmem populations predominated over S1-directed responses (p = 0.0147 and p = 0.0021 respectively) in late convalescence (Extended Data Fig 9B,D).

Polyclonal T cell responses to S comprise an array of immunodominant and subdominant epitopes; we therefore additionally tracked single CD4+ T cell epitopes in a subset of 9 donors (Extended Data Fig 10A). Strikingly, we observed substantial inter- and intra-individual variability in longitudinal epitope-specific responses (Extended Data Fig 10B,C); in some subjects, all epitope-specific responses tracked similarly while in others distinct epitope-specific responses would vary independently over time. In most, but not all, cases, peptide responses tracked similarly between the cTFH and Tmem populations (Extended Data Fig 10C). Overall, some degree of T cell immunity remains readily detectable in most subjects 4 months after infection, although longitudinal epitope-specific frequencies were markedly less predictable.

Deconvoluting the protective potential of the suite of concomitant immune responses elicited by SARS-CoV-2 infection is challenging. The general decline of serological immunity over time (Fig 4A) was similarly observed for most memory immune cell subsets except for IgG+ and IgA+ MBC populations (Fig 4B). Importantly, rates of immune decay are likely to stabilise over time to levels of homeostatic maintenance^37^, although this set point is not yet clear for SARS-CoV-2. Neutralising antibody is the most widely accepted protective correlate against a range of human respiratory viruses^38^. However, any relationship between *in vitro* neutralisation titres and *in vivo* protection for SARS-CoV-2 is unclear at present. We therefore developed a simulation model (see Methods) employing the estimated initial distributions of neutralisation titres and decay rates across subjects, to predict the time for titres to drop below a nominated cut-off of 1:40, selected based on the 1:40 hemagglutination inhibition titre (a surrogate for neutralisation activity) widely used as the 50% protective titre for influenza^39^. Notably, 43% of our cohort were already below this threshold in early convalescence, with 64% of subjects dropping below this threshold in late convalescence. Simulating a population of 1000 individuals, and running the model 1000 times, we find the median time for 50% of the population to drop below a titre of 1:40 was 74 days (Fig 4C; 95% confidence interval 46 to > 1000 days). Assuming early neutralisation titres predicted titres into late convalescence, our simulation also allows us to estimate how higher initial levels of neutralisation may affect the proportion of individuals maintaining titres above 1:40. We found that if aiming for a median of 50% of individuals with a titre above 1:40 at one year, initial neutralisation titres at about day 30 would need to be in the order of 2.1-fold higher than that observed in our convalescent cohort (95% CI = no increase to 16.9-fold increase required). It is important to emphasise that at present the in vitro neutralisation titre required and the additive contribution of other immune responses to protective immunity are unknown. In addition, our analysis assumes that immunity to vaccination decays at a similar rate to infection, and that the decay of neutralisation titre from day 70 to around 140 predicts immune decay over the first year. Despite the limitations inherent in these assumptions, this analysis provides an approach to estimating the target level of immune response necessary for effective vaccination.

Overall, we find that both neutralising and binding antibody responses decay as expected after recovery from COVID-19, assessed using polyclonal assays and at the level of single clonotypes. While incredibly durable protective antibody responses have been reported for other viral infections such as measles and smallpox^4^, our data suggests that SARS-CoV-2 is more likely to mirror immunity to endemic CCC, where serum antibody responses decline with a likelihood of increasing susceptibility to homologous virus within 1-2 years^5^. Neutralising antibody is a presumed but not yet proven correlate of immune protection for SARS-CoV-2. Assuming similar immune kinetics, our modelling suggests SARS-CoV-2 vaccines would likely need to elicit substantially more potent neutralising titres than infection to induce durable protection. Encouragingly, many early vaccine candidates have exceeded this metric when compared against sera from convalescent subjects in clinical trials reported to date^40,41^. Persistence of serum antibody is unlikely to be the sole determinant of long-lasting immunity, with anamnestic recall of stably maintained memory T and B cell populations likely reducing infection or disease. The magnitude, quality and protective potential of cellular responses against SARS-CoV-2 requires further definition. Although T cell memory in the blood contracts several months post infection, a rise in S-specific IgG+ memory B cells to a median level of ∼0.8% of all IgG+ memory B cells by 4 months suggests even mild-moderate COVID-19 induces substantial cellular immune memory.

## Materials and Methods

### Ethics Statement

The study protocols were approved by the University of Melbourne Human Research Ethics Committee (#2056689) and the Southern Adelaide Clinical Human Research Ethics Committee (#39.034), and all associated procedures were carried out in accordance with the approved guidelines. All participants provided written informed consent in accordance with the Declaration of Helsinki.

### Subject recruitment and sample collection

Subjects who had recovered from COVID-19 were recruited through contacts with the investigators and invited to provide serial blood samples. Subject characteristics of SARS-CoV-2 convalescent subjects are collated in Extended Data Fig 1. For all participants, whole blood was collected with sodium heparin anticoagulant. Plasma was collected and stored at −80°C, and PBMCs were isolated via Ficoll-Paque separation, cryopreserved in 10% DMSO/FCS and stored in liquid nitrogen.

### Microneutralisation Assay

SARS-CoV-2 isolate CoV/Australia/VIC01/2020^42^ was passaged in Vero cells and stored at −80C. Plasma was heat-inactivated at 56°C for 30 min. Plasma was serially-diluted 1:20 to 1:10240 before addition of 100 TCID_50_ of SARS-CoV-2 in MEM/0.5% BSA and incubation at room temperature for 1 hour. Residual virus infectivity in the plasma/virus mixtures was assessed in quadruplicate wells of Vero collected and stored at −80ºC, and PBMCs were isolated cells incubated in serum-free media containing 1 µg/ml TPCK trypsin at 37°C/5% CO2; viral cytopathic effect was read on day 5. The neutralising antibody titre is calculated using the Reed/Muench method as previously described^43,44^. All samples were assessed in two independent microneutralisation assays.

### Expression of SARS-CoV-2 proteins

A set of proteins was generated for serological and flow cytometric assays. The ectodomain of SARS-CoV-2 (isolate WHU1;residues 1 – 1208) was synthesised with furin cleavage site removed and P986/987 stabilisation mutations^45^, a C-terminal T4 trimerisation domain, Avitag and His-tag, expressed in Expi293 cells and purified by Ni-NTA affinity and size-exclusion chromatography using a Superose 6 16/70 column (GE Healthcare). SARS-CoV S was biotinylated using Bir-A (Avidity). The SARS-CoV-2 RBD^46^ with a C-terminal His-tag (residues 319-541; kindly provided by Florian Krammer) was similarly expressed and purified.

### SARS-CoV-2 bead-based multiplex assay

The isotypes and subclasses of SARS-CoV-2 specific antibodies were detected as previously described^25^. Briefly, a panel of SARS-CoV-2 antigens including trimeric S, S1 (Sino Biological), S2 (ACROBiosystems), NP (ACROBiosystems,) and RBD^46^ were coupled to magnetic COOH- bioplex beads (Biorad) using a two-step carbodiimide coupling reaction. 20μl of bead mixture containing 1000 beads per region and 20μl of 1:200 diluted plasma were added per well. SARS-CoV-2- specific antibodies were detected using phycoerythrin (PE)-conjugated mouse anti-human pan-IgG, IgG1, IgG2, IgG3, IgA1 or IgA2 (Southern Biotech) at 1.3μg/ml, 25μl per well. For the detection of IgM, biotinylated mouse anti-human IgM (mAb MT22; MabTech) was added at 1.3μg/ml, 25μl per well followed by streptavidin-PE (SA-PE; Thermo Fisher) at 1μg/ml. Plates were acquired by a FLEXMAP 3D (Luminex). Median fluorescence intensity (MFI) for each isotype/subclass detector was assessed. Background subtraction was conducted, removing background of blank (buffer only) wells. Multiplex assays were repeated twice as two independent experiments.

### RBD-ACE2 binding inhibition multiplex bead-based assay

RBD protein was coupled to bioplex beads (Biorad) as described above. 20μl of RBD multiplex bead suspension containing 500 beads per well, 20μl of biotinylated Avitag-ACE2 (kindly provided by Dale Godfrey and Nicholas Gherardin), final concentration of 12.5μg/ml per well, along with 1:100 dilution of each subject’s plasma were added to 384 well plates. Plates were covered and incubated at room temperature (RT) whilst shaking for 2 hours, and then washed twice with PBS containing 0.05% Tween20 (PBST). Biotinylated Avitag-ACE2 was detected using 40μl per well of SA-PE at 4μg/ml, incubated with shaking for 1 hour at RT. 10μl of PE-Biotin amplifier (Thermo Fisher) at 10μg/ml was added and incubated for 1 hour with shaking at RT. Plates were washed and acquired on a FLEXMAP 3D (Luminex). Anti-SARS-CoV-2 RBD neutralising human IgG1 antibody (ACROBiosystems, USA) was included as a positive control, in addition to COVID-19 negative plasma and buffer only negative controls. The MFI of bound ACE2 was measured after background subtraction of no ACE2 controls. Maximal ACE2 binding MFI was determined by buffer only controls. % ACE2 binding inhibition was calculated as 100% – (% ACE2 binding MFI per sample/ Maximal ACE2 binding). RBD-ACE2 binding inhibition multiplex assays were repeated independently twice.

### Flow cytometric detection of S- and RBD-specific memory B cells

Probes for delineating SARS-CoV-2 S-specific B cells within cryopreserved human PBMC were generated by sequential addition of streptavidin-PE (Thermofisher) to trimeric S protein biotinylated using recombinant Bir-A (Avidity). SARS-CoV-2 RBD protein was directly labelled to APC using an APC Conjugation Lightning-link kit (Abcam). Cells were stained with Aqua viability dye (Thermofisher). Monoclonal antibodies for surface staining included: CD19-ECD (J3-119) (Beckman Coulter), CD20 Alexa700 (2H7), IgM-BUV395 (G20-127), CD21-BUV737 (B-ly4), IgDCy7PE (IA6-2), IgG-BV786 (G18-145) (BD), CD14-BV510 (M5E2), CD3-BV510 (OKT3), CD8a-BV510 (RPA-T8), CD16-BV510 (3G8), CD10-BV510 (HI10a), CD27-BV605 (O323) (Biolegend), IgA-Vio450 (clone) (Miltenyi). Cells were washed, fixed with 1% formaldehyde (Polysciences) and acquired on a BD LSR Fortessa or BD Aria II.

### Flow cytometric detection of antigen-specific cTFH, memory CD4+ T cells and memory CD8 T cells

Cryopreserved human PBMC were thawed and rested for four hours at 37°C. Cells were cultured in 96-well plates at 1×10^6^ cells/well and stimulated for 20 hours with 2μg/peptide/mL of peptide pools (15mer, overlapping by 11) covering the S1 or S2 domains of SARS-CoV-2. Selected donors were also stimulated with SEB (1μg/mL) as a positive control, or individual peptides at 2ug/mL: NCTFEYVSQPFLMDL (S1 epitope; previously described in^47^); LPIGINITRFQTLLA (S1 epitope); GWTFGAGAALQIPFA (S2 epitope); ALQIPFAMQMAYRFN (S2 epitope); LLQYGSFCTQLNRAL (S2 epitope;^19,47^); QALNTLVKQLSSNFG (S2 epitope). Following stimulation, cells were washed, stained with Live/dead Blue viability dye (ThermoFisher), and a cocktail of monoclonal antibodies: CD27 BUV737 (L128), CD45RA PeCy7 (HI100), CD20 BUV805 (2H7), (BD Biosciences), CD3 BV510 (SK7), CD4 BV605 (RPA-T4), CD8 BV650 (RPA-T8), CD25 APC (BC96), OX-40 PerCP-Cy5.5 (ACT35), CD69 FITC (FN50), CD137 BV421 (4B4-1) (Biolegend), and CXCR5 PE (MU5UBEE, ThermoFisher). Cells were washed, fixed with 1% formaldehyde and acquired on a BD LSR Fortessa using BD FACS Diva.

#### Mass spectrometry (MS)-based quantitative proteomics of serum anti-S1 antibodies

The workflow for anti-S1 proteomic profiling is shown in Extended Data Figure 4A. Briefly, antibodies against SARS-CoV-2 S1 spike protein were affinity-purified from convalescent plasma of COVID-19 subjects at different time points using S1 protein-coupled magnetic beads (Acrobiosystems). IgG heavy chains were isolated after reduced SDS-PAGE and digested with trypsin and chymotrypsin to generate peptides for LC-MS/MS using a Thermo Scientific Orbitrap Exploris 480 mass spectrometer coupled to an Ultimate 3000 UHPLC (Dionex). De novo sequencing data analysis was performed by Peaks studio X-plus software (Bioinformatics Solution). Peptide sequences were referenced against recovered heavy chain immunoglobulin sequences generated from single sorted S-specific memory B cells^19^ to identify matched CDR-H3 peptides. Anti-S1 clonotypic antibody expression levels were monitored by parallel reaction monitoring (PRM) as described previously^28,48^. Fragment ion extracted ion chromatograms (XICs) per CDR-H3 peptide were visualized in Skyline version 20.1.0.155 (University of Washington) and inspected manually to ensure correct assignments. The annotated spectra of individual peptides and their corresponding XICs are shown in Supplementary Information.

#### Estimating the decay rates

We sought to predict the response variable (y_ij_ for patient i at timepoint j) as a function of days post symptom onset, assay replicate (as a binary categorical variable) and a random effect for each individual (both in in intercept and slope). The dependency of the response variables on days post symptom onset can be modelled by using one or two decay slopes. The model can be written as below: *y_ij_* = *β*_0_ + *b*_0*i*_ + *β*_1_*R_ij_* + *β*_2_*t_ij_* + *b*_2*i*_*t_ij_* – for a model with a single slope; and *y_ij_* = *β*_0_ + *b*_0*i*_ + *β*_1_*R_ij_* + *β*_2_*t_ij_* + *b*_2*i*_*t_ij_* + *β*_3_*s_ij_* + *b*_3*i*_*s_ij_* – for a model with two different slopes, in which:

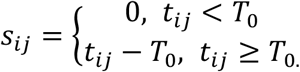

The parameter *β*_0_ is a constant (intercept), and *b*_0*i*_ is a patient-specific adjustment to the overall intercept. The slope parameter *β*_2_ is a fixed effect to capture the decay slope before *T*_0_; which also has a subject-specific random effect *b*_2*i*_. To fit a model with two different decay rates, an extra parameter *β*_3_ (with a subject-specific random effect *b*_3*i*_) was added to represent the difference between the two slopes. Assay variability between replicates was modelled as a single fixed effect *β*_1_, in which we coded the replicate as a binary categorical variable *R_ij_*.

The response variables obtained were highly variable, containing zeros where the value was below the limit of detection and contrasted with samples where very high levels were observed. Thus, we performed log transformations of the non-zero data to help normalize variability and censored every value less than 40 for the microneutralisation data; every value less than 0.01 for the T cell and B cell data; and every negative value for the multiplex data. More specifically, a mixed-effect regression method that allows for censoring at the limit of detection was used to estimate the parameters in the model. This was done by using *lmec* library in *R*, using the ML algorithm to fit for the fixed effects^49^. We also tested if the decay of serological response variables was fitted better by a single or two different decay slopes (likelihood ratio test – based on the likelihood value and the difference in the number of parameters). 95% CI for the fixed effect parameters was calculated based on the standard error estimates, which can be obtained directly by using the *varFix* function from *lmec* library. These analyses were carried out in *R* version 4.0.2.

### Simulating the decay of serological neutralisation activity

To understand the decay in serum neutralisation we employed a simulation approach using the parameters estimated from our mixed-effect censoring regression model of decay. The fixed effect estimates averaged the intercept across experimental replicates (β_0_ + β_1/2_ from equations above) and random effects were randomly selected from a multivariate normal distribution with covariance matrix taken from the mixed-effect regression with censoring lmec object. The residual error standard deviation for simulated data every 10 days was taken from the lmec object. The confidence interval for the percentage of subjects with a neutralisation titre above 1:40 was estimated empirically with the percentile method by repeating the simulation 1000 times, where for each replicate the fixed effects were drawn from a normal distribution based on their standard error (as well as randomly selected random effects). To estimate the fold increase in initial neutralisation titre required to achieve > 50% of individuals with a titre above 40 at 1 year we assumed that the rate of decay was constant from day 70 onwards and projected forward the expected titres in the simulated populations. The median and confidence intervals for the proportion of individuals with titre > 40 were calculated from these 1000 simulated populations.

### Statistical Analyses

Associations between neutralisation, inhibition of ACE2 binding and antibody binding were assessed using both Spearman correlation, and multiple regression (R version 4.0.2). The geometric mean of replicate neutralisation measurements and the arithmetic mean of replicate measurements in other assays were used in the correlation and regression analyses for other measurements. Neutralisation titres below the limit of detection (a titre of 20) were assigned the arbitrary value 10 prior to calculating the geometric mean for the purposes of the Spearman correlation, where rank and not magnitude of the measurements is important. For the multiple regression analysis values below the limit of detection were set at the detection threshold and censoring regression was performed using the function *censReg (*from the *censReg* library^50^) to determine which measurements during early convalescence were significant predictors of neutralisation titre during late convalescence. Comparison of B and T cell frequencies at first and final sampling was performed using Wilcoxon Rank Sum test in GraphPad Prism 8. All statistical tests used were two sided.

## Data Availability

All data are available from the corresponding author upon reasonable request.

## Data Availability

All data are available from the corresponding author upon reasonable request.

## Code Availability

All data analysis code can be made available on request.

## Authors’ contributions

A.K.W, J.A.J., J.J.W, H.-X.T., T.E.S., D.L.G., D.S.K., D.C., T.P.G, A.W.C., M.P.D., and S.J.K. designed the study and experiments. A.K.W., J.A.J., J.J.W., K.J.S, A.R., H.-X.T., W.S.L., K.M.W., H.G.K., R.E., S.K.D, H.E.K, F.L.M., T.E.S., and K.S. performed experiments. A.K.W., J.A.J., J.J.W, H.-X.T., W.S.L., A.R., D.S.K., T.E.S., D.C., K.S., M.P.D., S.J.K. analysed the experimental data. A.K.W, J.A.J., J.J.W, K.J.S, T.E.S., D.S.K., D.C., M.P.D., and S.J.K. wrote the manuscript. All authors reviewed the manuscript.

## Acknowledgements

We thank the generous participation of the trial subjects for providing samples. We thank E. Haycroft, E. Lopez, C. Nelson and T. Amarasena and C. Batten (University of Melbourne) for excellent technical assistance. The SARS-CoV-2 RBD expression plasmids were kindly provided by F. Krammer (Icahn School of Medicine at Mt Sinai). Recombinant human ACE2 was kindly provided by N. Gherardin and D. Godfrey (University of Melbourne). We acknowledge the Melbourne Cytometry Platform (Melbourne Brain Centre node) for provision of flow cytometry services. We thank T. Chataway and A. Colella (Flinders Proteomics Facility) for technical support with quantitative proteomics. This study was supported by the Victorian Government, an Australian government Medical Research Future Fund award GNT2002073 (SJK, MPD, and AKW), the ARC Centre of Excellence in Convergent Bio-Nano Science and Technology (SJK), an NHMRC program grant APP1149990 (SJK and MPD), NHMRC project grant GNT1162760 (AKW), an NHMRC-EU collaborative award APP1115828 (SJK and MPD), the European Union Horizon 2020 Research and Innovation Programme under grant agreement 681137 (SJK), Emergent Ventures Fast Grants (AWC), the Jack Ma Foundation (KS) and the A2 Milk Company (KS). JAJ, DSK, and SJK are supported by NHMRC fellowships. JJW is supported by Flinders University DVCR Fellowship and Flinders Health & Medical Research Institute COVID-19 Research Grant. AKW, KS, DC and MPD are supported by NHMRC Investigator grants. The Melbourne WHO Collaborating Centre for Reference and Research on Influenza is supported by the Australian Government Department of Health.

---

**Supplementary Fig. S1:**
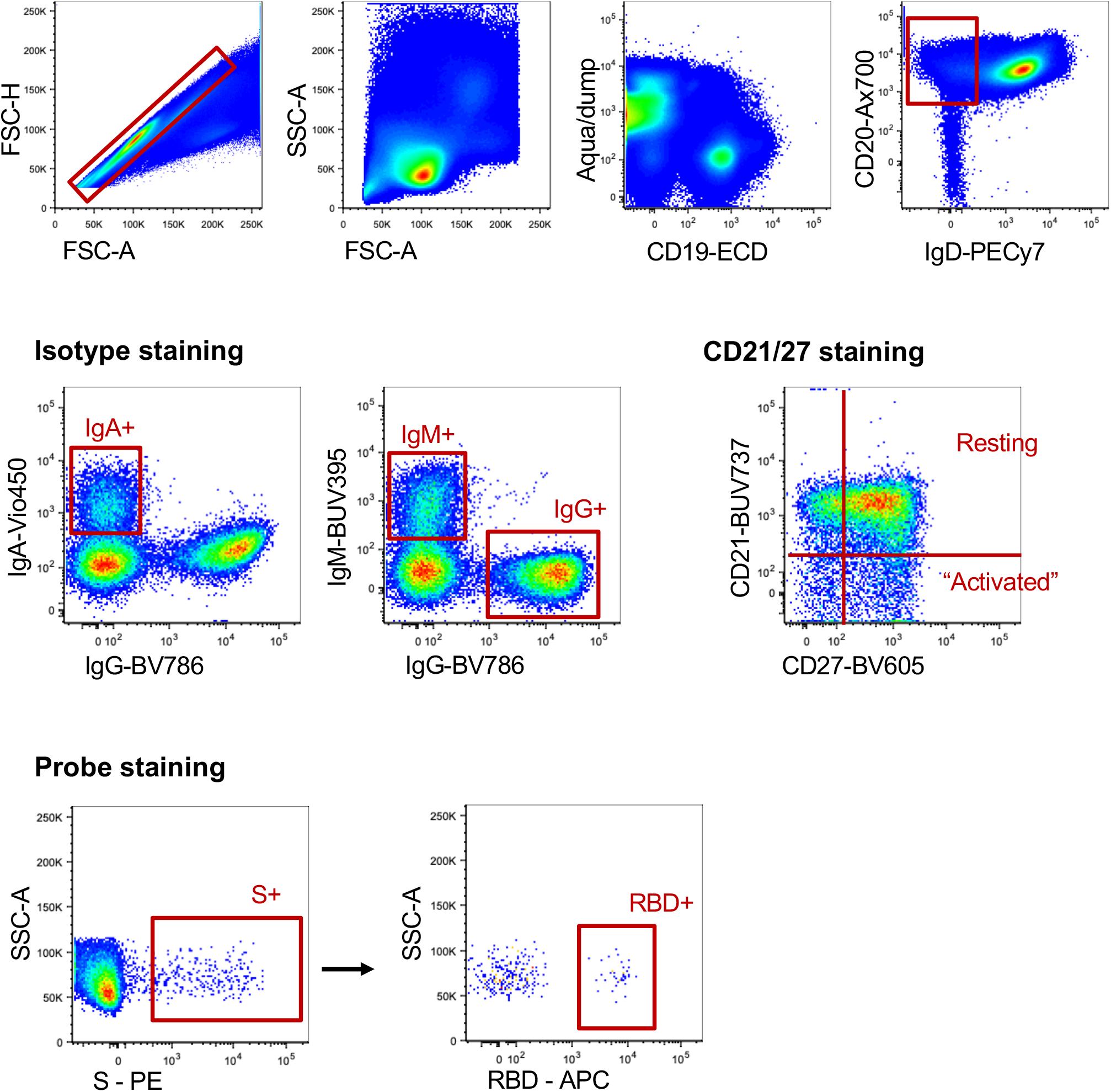
Gating strategy for resolving antigen-specific B cells and isotypes. After doublet exclusion (FSC-A vs FSC-H) and lymphocyte gating (FSC-A vs SSC-A), live +IgD-CD20+ B cells were gated based on surface immunoglobulin expression (IgM, IgA). Binding to SARS-CoV-2 spike (S) and/or SARS-CoV-2 RBD probes was for each population. Memory B cell phenotypes were identified by CD21 and co-staining.

**Supplementary Fig. S2:**
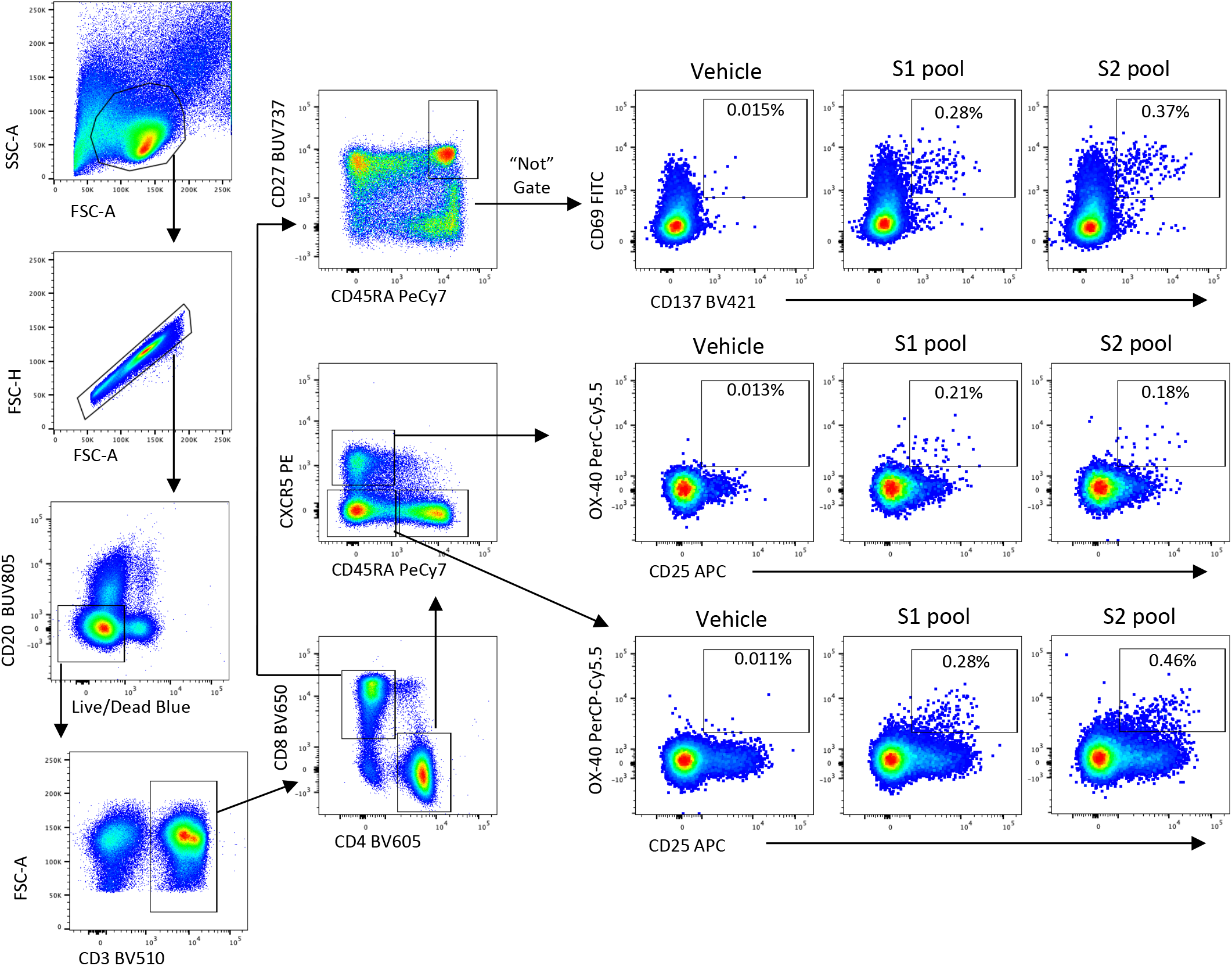
Gating strategy for quantifying antigen-specific T cells. Lymphocytes were identified by FSC/SSC, followed by doublet exclusion (FSC-A vs FSCH), and exclusion of dead or CD20+ cells. After gating on CD3, single positive CD4 or CD8 subsets were identified. CD8 Tmem were gated as non-naïve (CD27+CD45RA+) and assessed for co-expression of CD69 and CD137 following stimulation. CD4 T were gated as cTFH (CXCR5+CD45RA-) or Tmem (CXCR5-CD45RA-), and assessed –expression of OX-40 and CD25 following stimulation.

**Supplementary Fig. S3:**
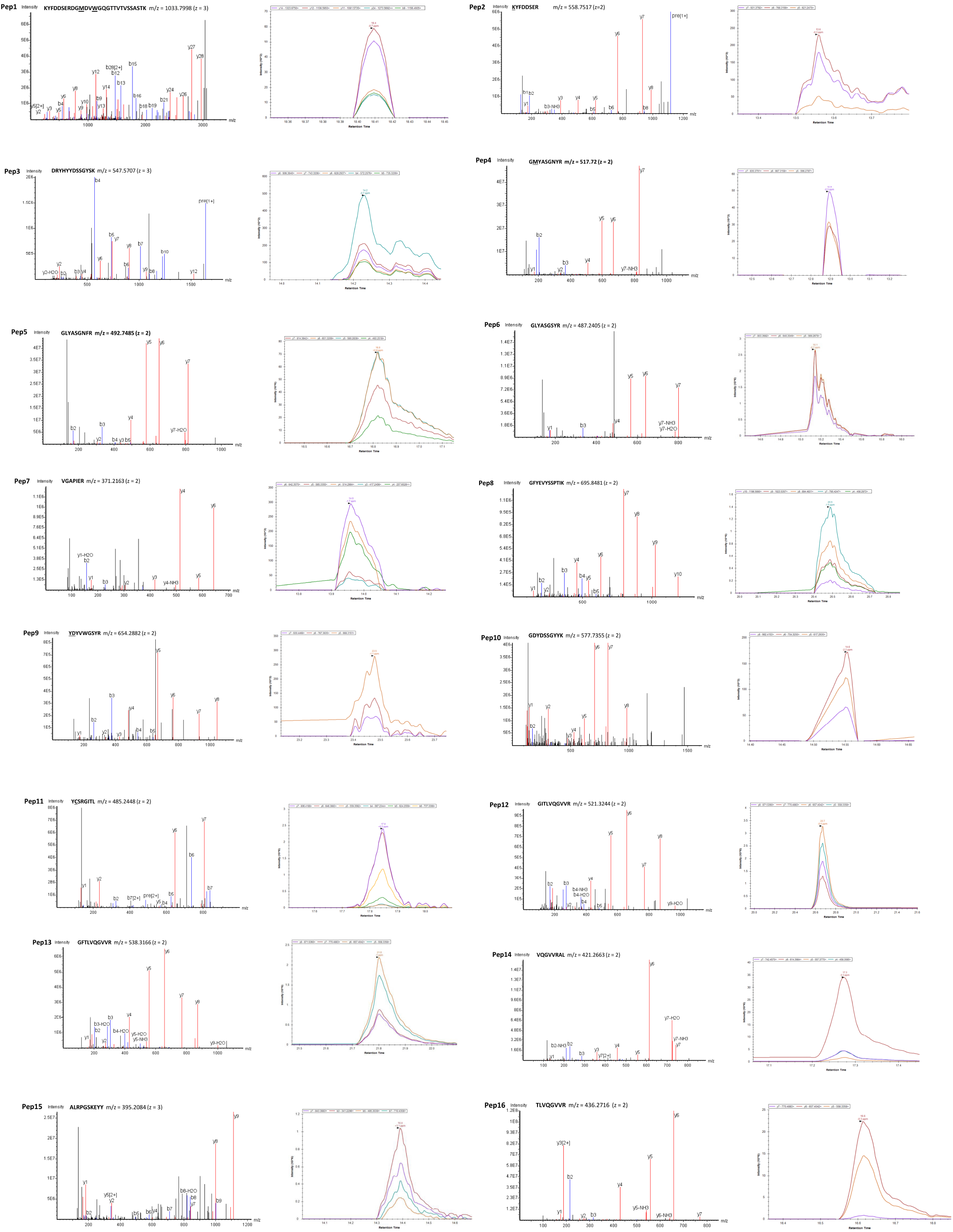
Representative annotated MS/MS spectra (left panel) and their corresponding extracted ion chromatograms (XICs; right panel). The peptides used in PRM analyses are the matched clonotypic CDR-H3 peptides (pep1–16). Underlined amino acid indicates a post translational modification: <u>M</u> (oxidised methionine), <u>W</u> (oxidised tryptophan), <u>K</u> (carbamidomethylated lysine), <u>D</u> (carbamidomethylated aspartate), <u>C</u> (carbamidomethylated cysteine), and <u>Y</u> (acetylated tyrosine). The sequences, m/z and z of each individual peptides are hown on the top of their annotated MS/MS spectra. Matched b ions are indicated in blue and y ions are in red. m = mass, z = charge.

## Extended Data Figures

**Extended Data figure 1.**
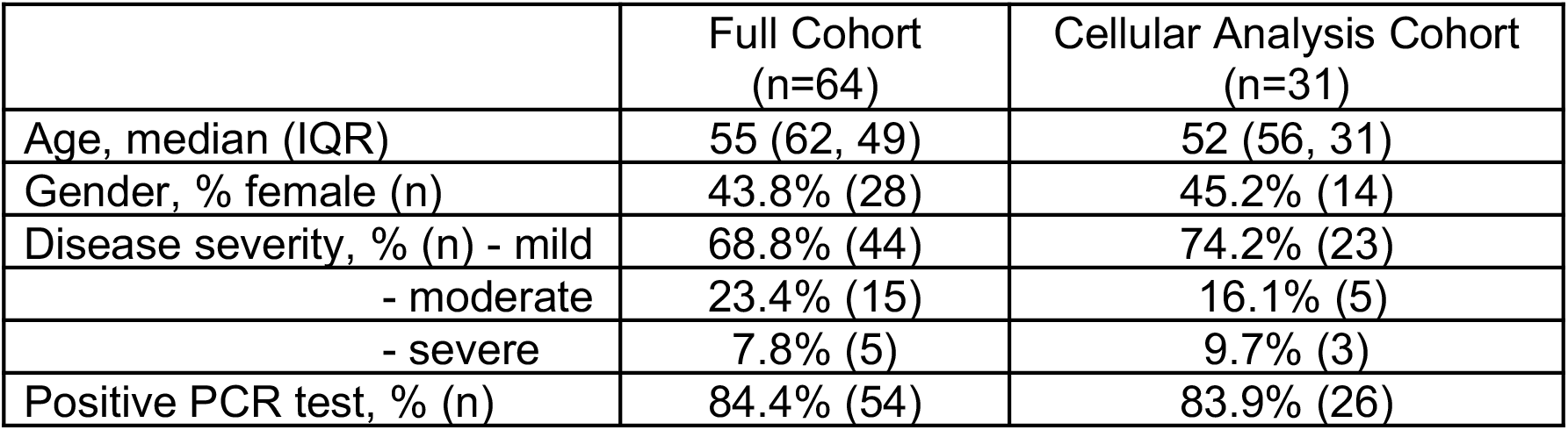
Demographic and clinical characteristics of the convalescent COVID-19 cohort. The best-fit model and half-lives are shown for the fitting of the decay of antibody binding to different SARS-CoV-2 antigens (n = 64 subjects). Two-phase decay is indicated by red (before day 70) and blue (after day 70) shaded areas. No shading indicates where single-phase decay provided the best fit.

**Extended Data figure 2:**
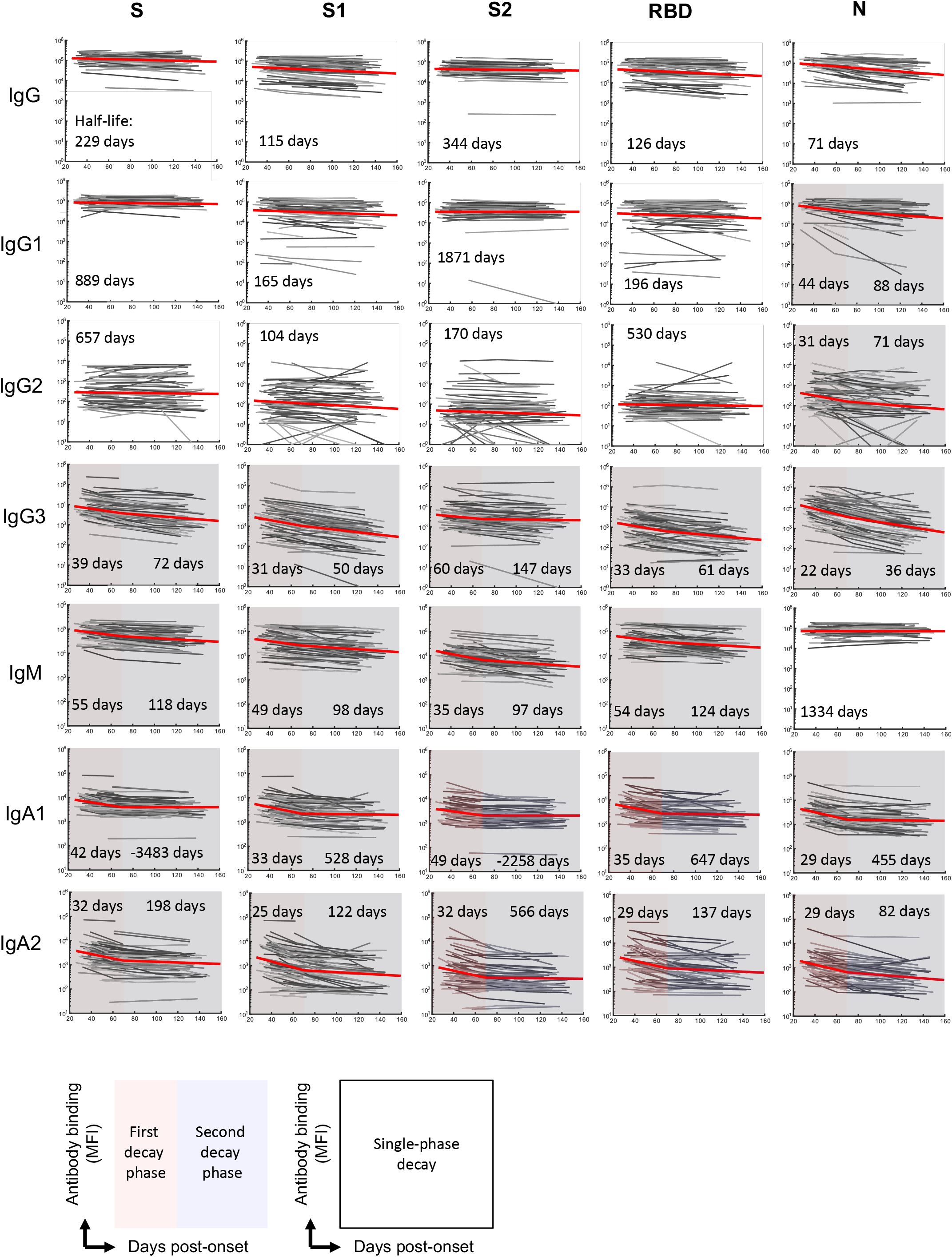
Fitting of the decline in antibody binding across different immunoglobulin isotypes. The best-fit model and half-lives are shown for the fitting of the decay of antibody binding to different SARS-CoV-2 antigens (n = 64 subjects). Two-phase decay is indicated by red (before day 70) and blue (after day 70) shaded areas. No shading indicates where single-phase decay provided the best fit.

**Extended Data Figure 3:**
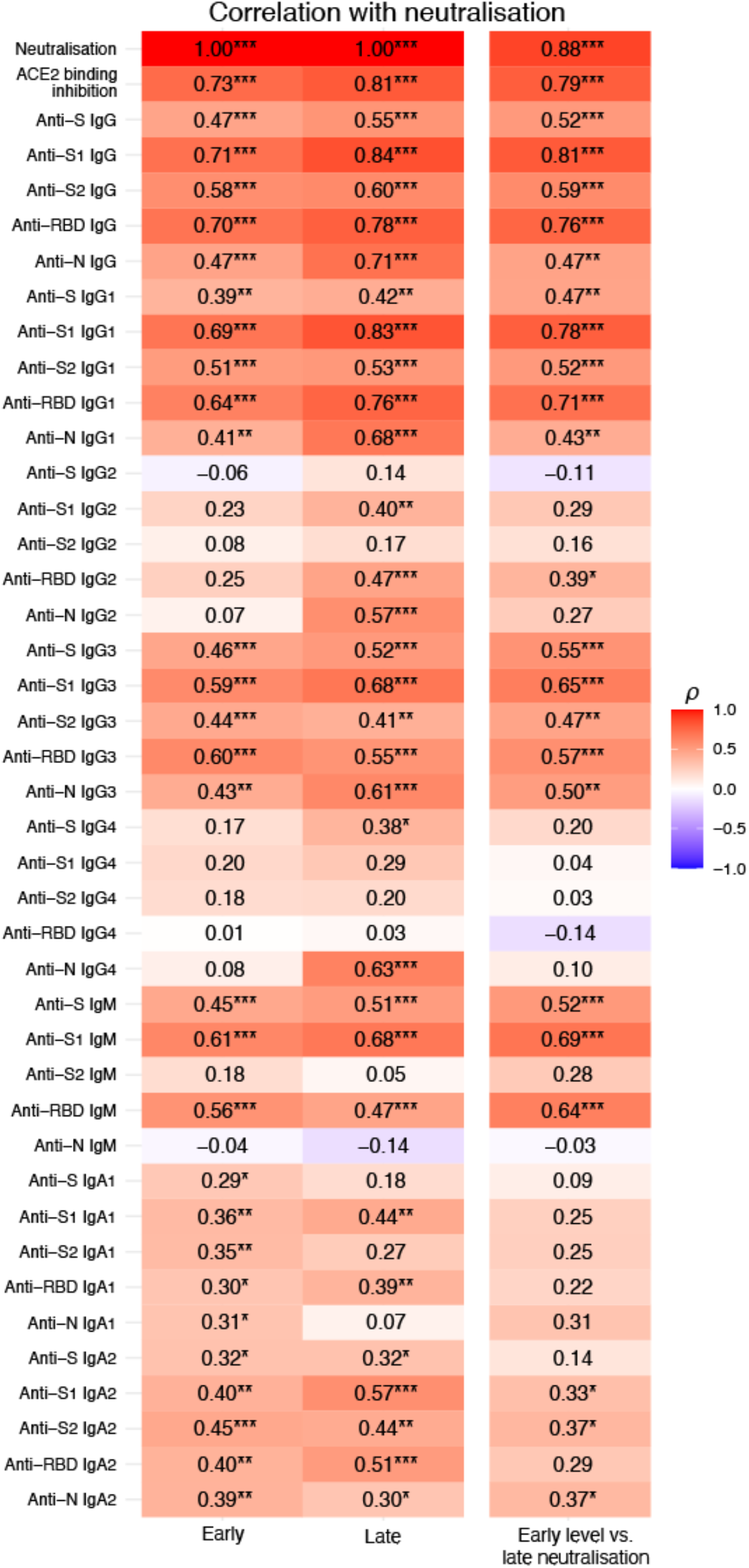
Correlation of antibody binding and ACE2 inhibition with neutralisation. A heat-map of Spearman correlations between neutralisation titre and the serological measurements of antibody binding (by isotype and antigen). Correlations were assessed in early (≤50 days, left column n = 54 subjects) and late (≥100 days, right middle column, n = 47 subjects) convalescence in all subjects were data was available. The association between early antibody binding and late neutralisation is also shown (right column, n = 47 subjects). All correlations are Spearman correlations. *P≤0.05, **P≤0.01, ***P≤0.001.

**Extended Data Figure 4:**
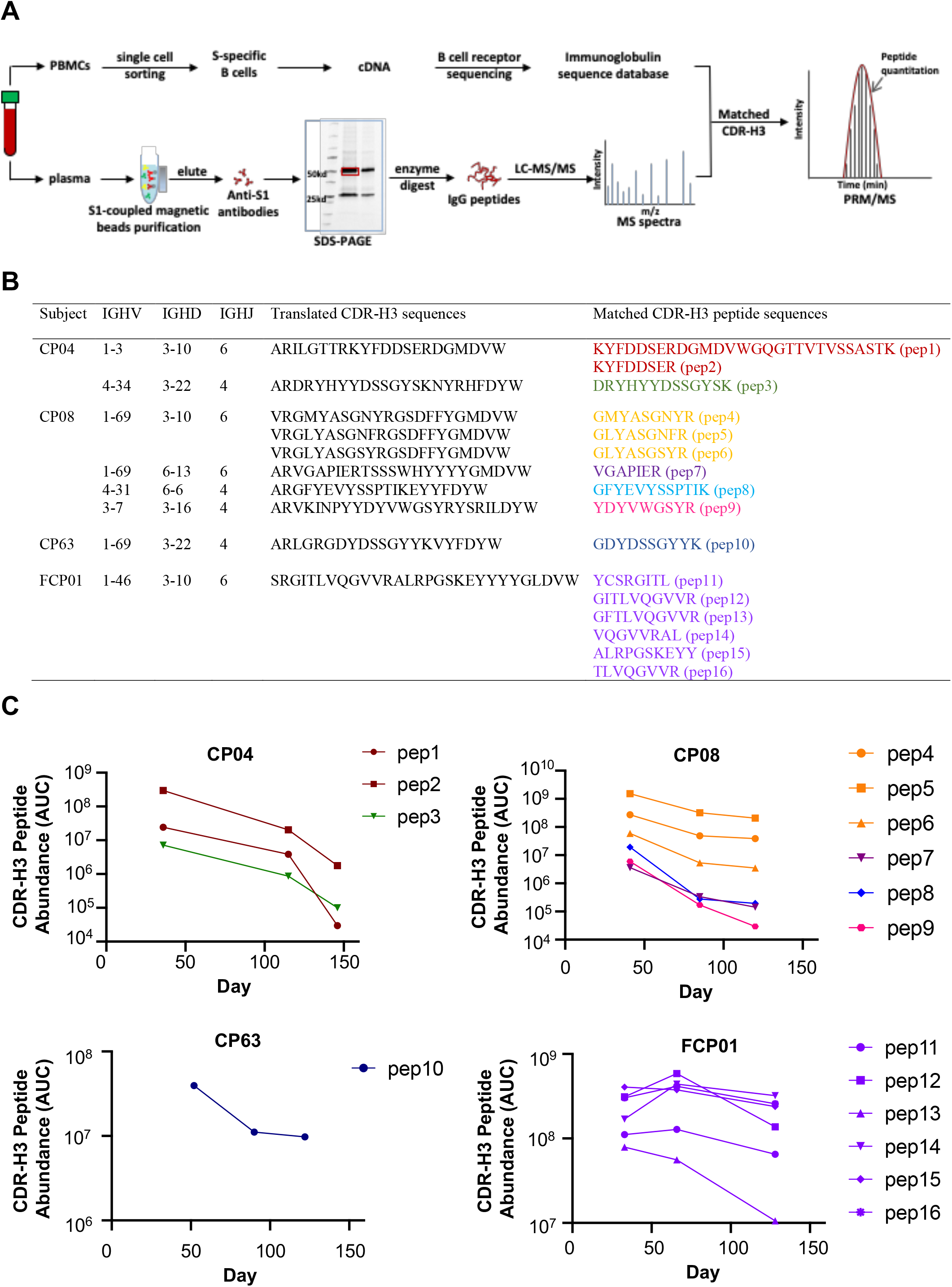
MS-based quantification of immunoprecipitated S1-specific clonotypic antibodies. **(A)** Combined B cell receptor sequencing and proteomics platform enables identification and quantification of circulating anti-S1 antibodies. S1-specific IgG was purified from plasma of SARS-CoV-2 convalescent subjects using antigen-coupled magnetic beads and heavy chains subject to LC-MC/MS. Peptide spectra are searched against B-cell receptor sequencers recovered from single sorted S-specific memory B cells from the same individuals to identify clonotypes based upon CDR-H3 amino acid sequence. Clonotype specific peptides are then used as barcodes for relative quantitative parallel reaction monitoring (PRM) for tracking in longitudinal plasma samples. Targeted peptides are monitored during elution from HPLC and individual peptides quantified based on abundance chromatography curves. **(B)** Clonotypes identified based on matched CDR-H3 sequences from S1-specific plasma IgG and B cell receptor sequences from SARS-CoV-2 convalescent subjects (n = 4). **(C)** Longitudinal changes in the relative plasma abundance of anti-S1 clonotypes within four convalescent subjects over time. The quantity of each reference peptide is expressed as area under the curve (AUC) derived from extracted ion chromatography.

**Extended Data Figure 5:**
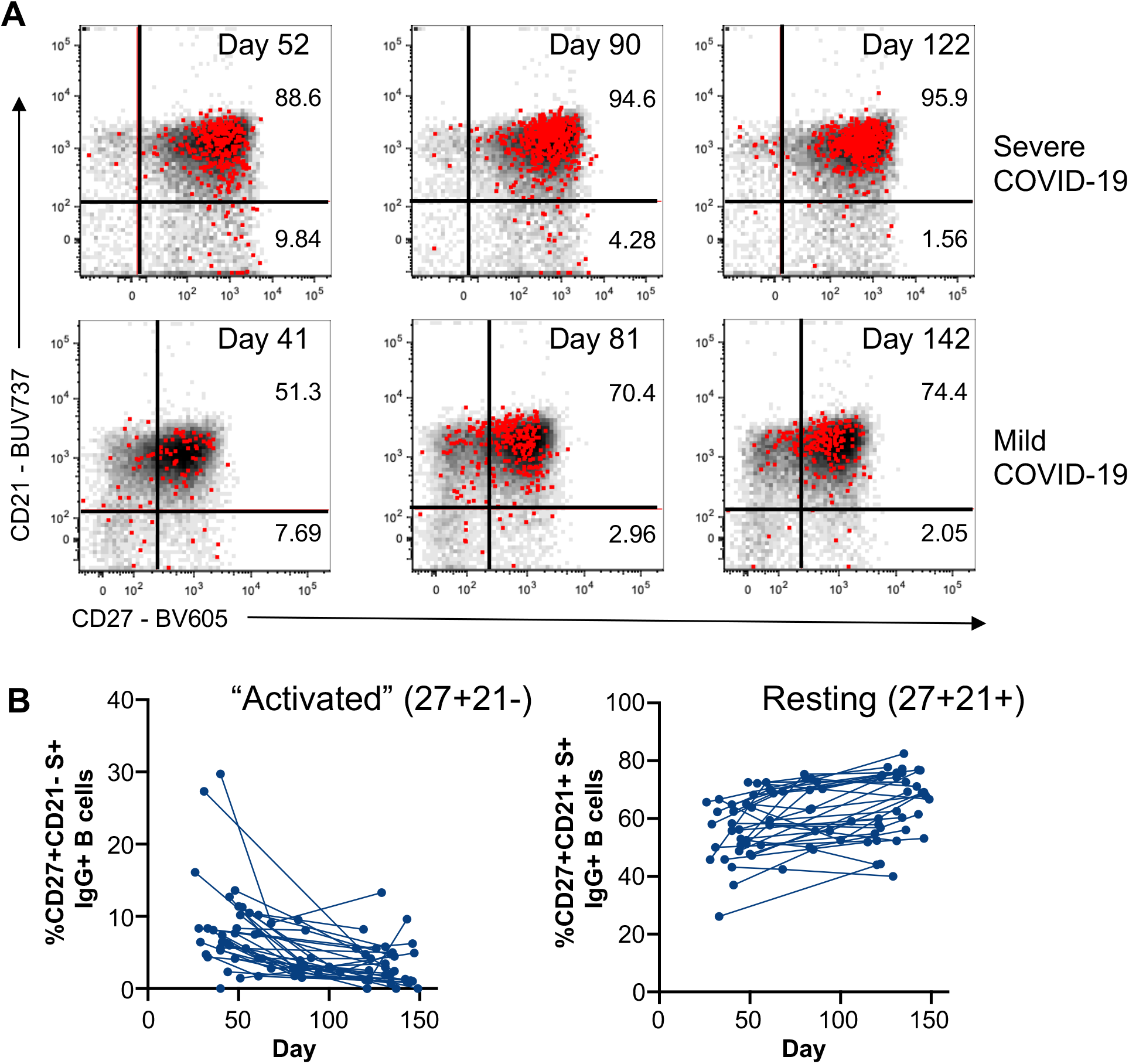
Activation status of S-specific IgG+ memory B cells. **(A)** Memory B cell phenotypes identified by CD21 and CD27 co-staining of S+ CD19+CD20+IgD-IgG+ B cells (red) overlaid onto parental CD19+CD20+IgD-IgG+ B cells (black) and **(B)** the corresponding frequencies of “activated” (CD27+CD21-) or resting (CD27+CD21+) in in PBMC samples were assessed longitudinally (n = 31 subjects).

**Extended Data Figure 6:**
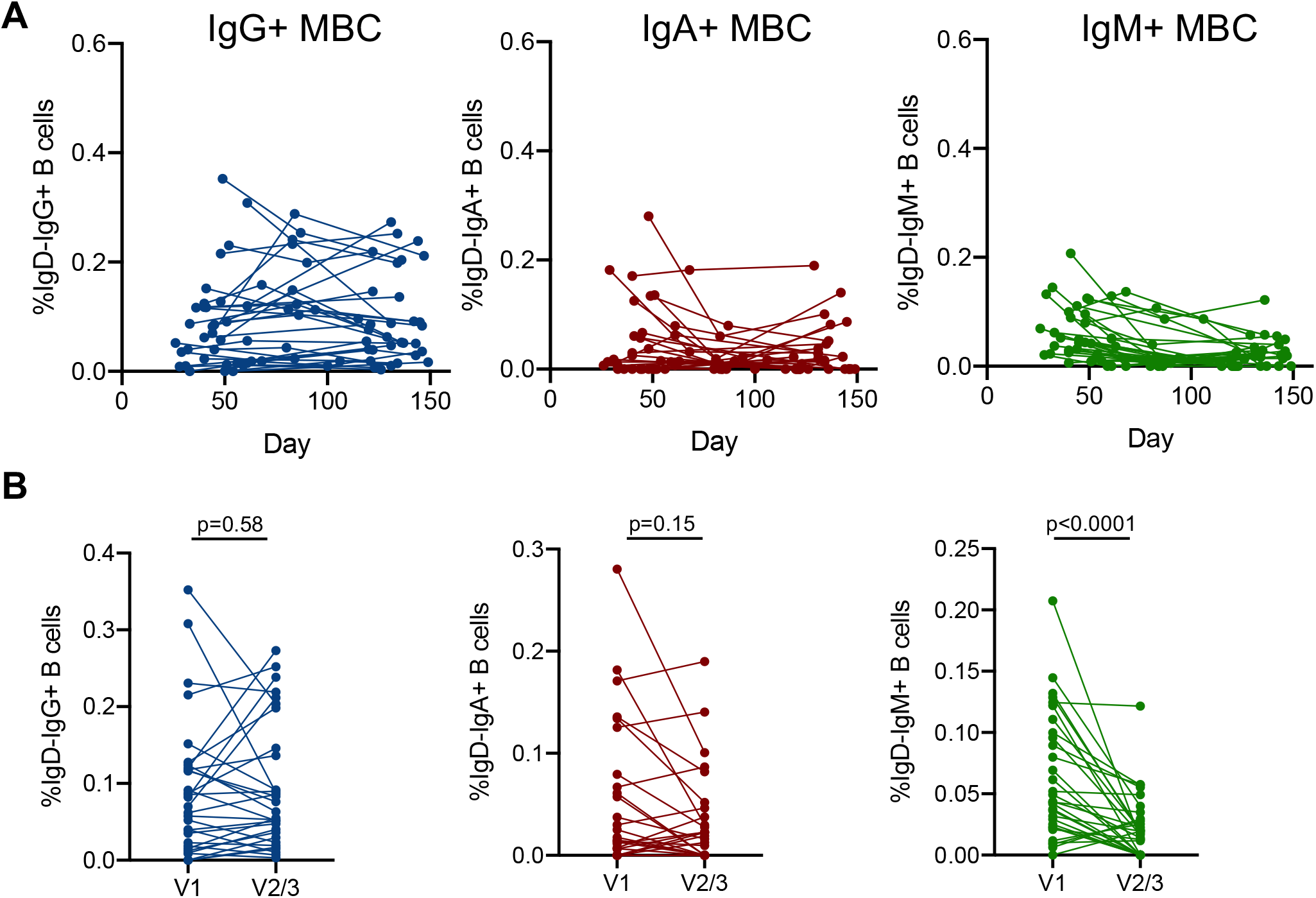
RBD-specific memory B cell dynamics. **(A)** Frequencies of RBD-specific IgG+, IgA+ or IgM+ memory B cells as a proportion of CD19+CD20+IgD- B cells in PBMC samples were assessed longitudinally. **(B)** Comparison of RBD-specific IgG+, IgA+ or IgM+ memory B cell frequencies at the earliest and latest timepoint available for each individual (n = 31). Statistics assessed by two-tailed Wilcoxon test.

**Extended Data Figure 7:**
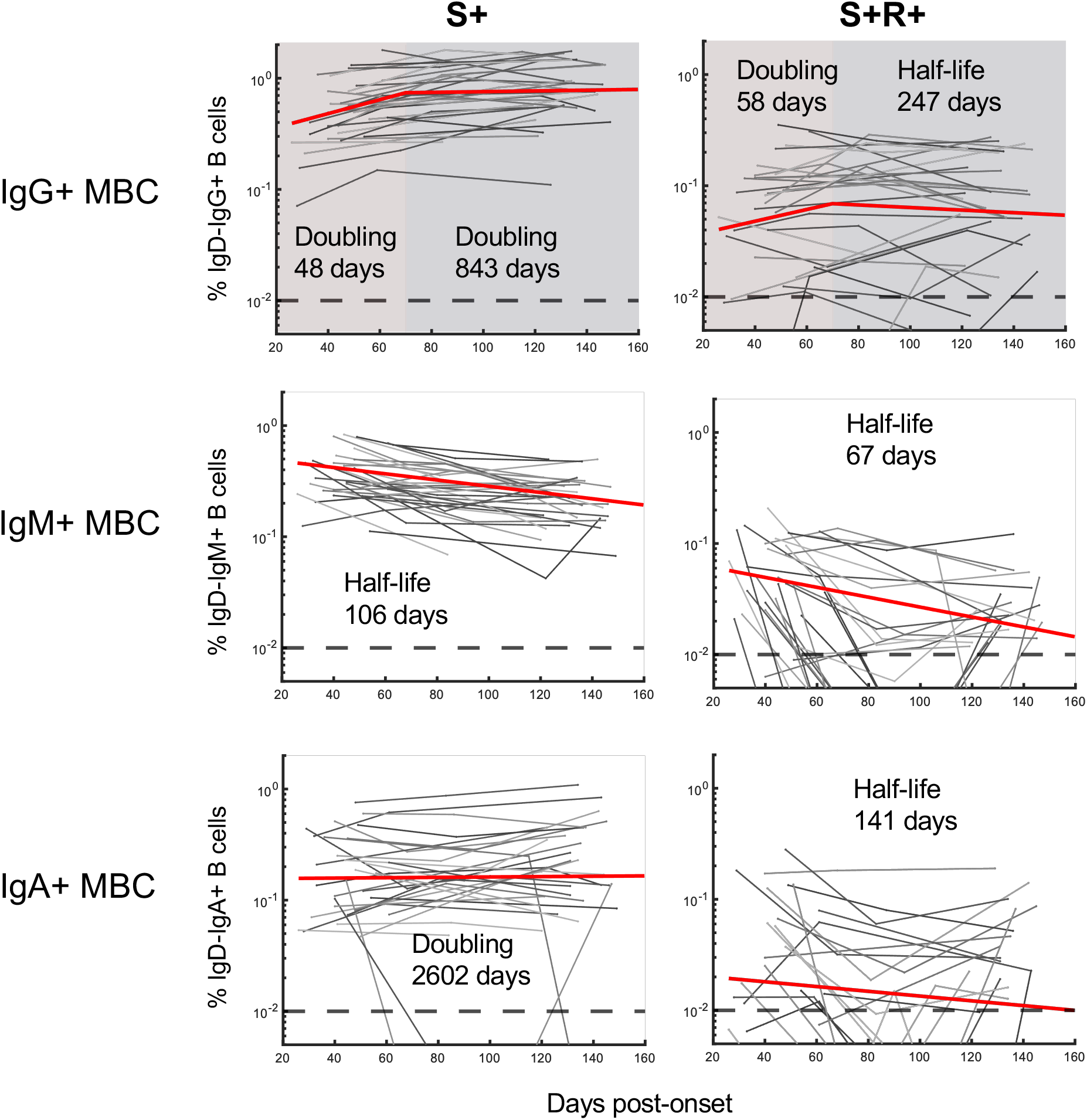
Fitting of the kinetics of S- and RBD-specific memory B cell responses over time. The best-fit half-lives are shown for the fitting of the growth and/or decay of S- or RBD-specific memory B cells (n = 31 subjects). Two-phase decay is indicated by red (before day 70) and blue (after day 70) shaded areas. No shading indicates where a single-phase decay model was used to fit the data.

**Extended Data Figure 8:**
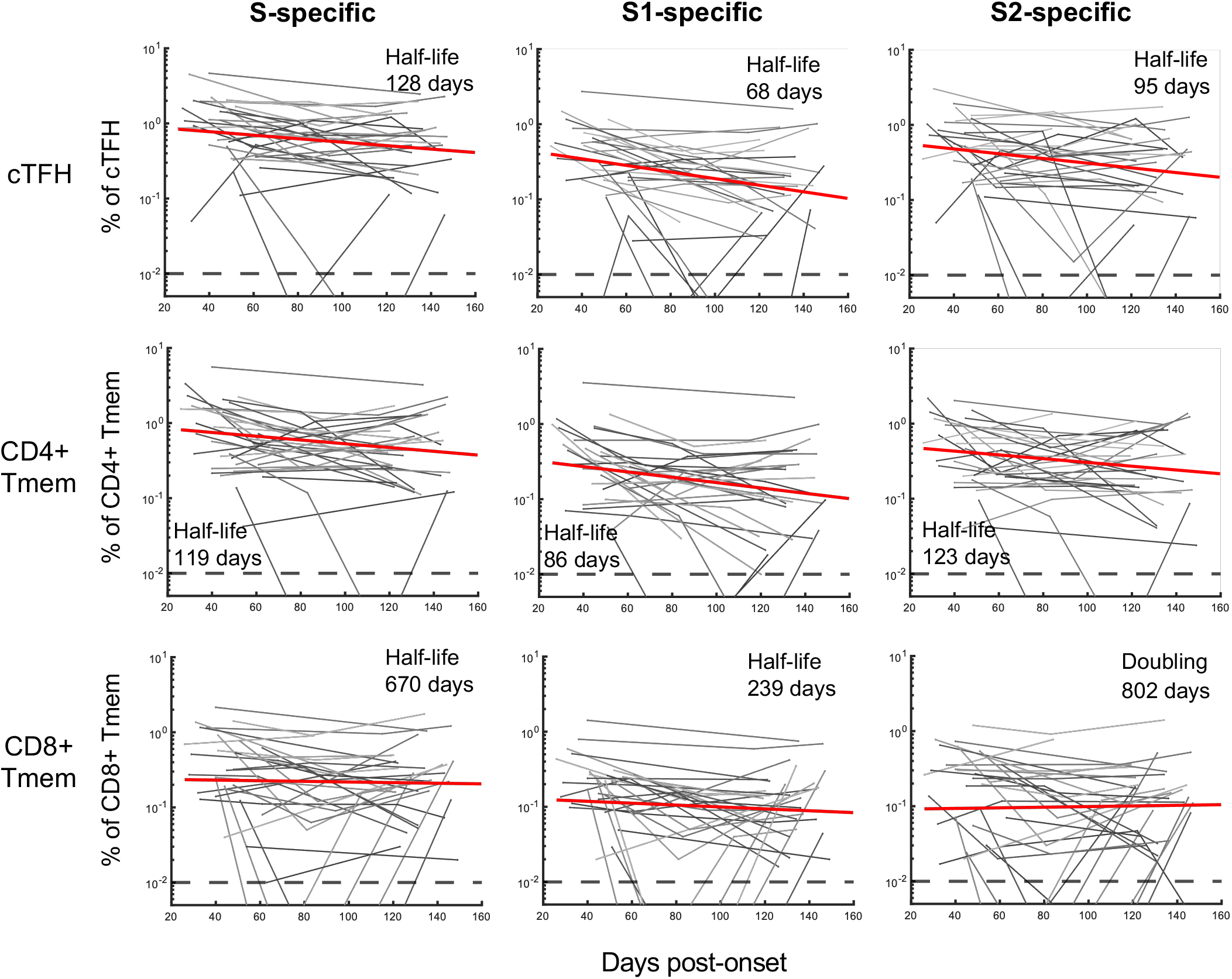
Fitting of the decline in SARS-CoV-2-specific T cells over time. The best-fit half-lives are shown for the fitting of the decay of cTFH, CD4+ Tmem and CD8+ Tmem specific to total S (S1+S2 responses combined), S1 or S2 peptide pools (n = 31 subjects). In all cases decay was fit with a single-phase decay model with the half-lives shown.

**Extended Data Figure 9:**
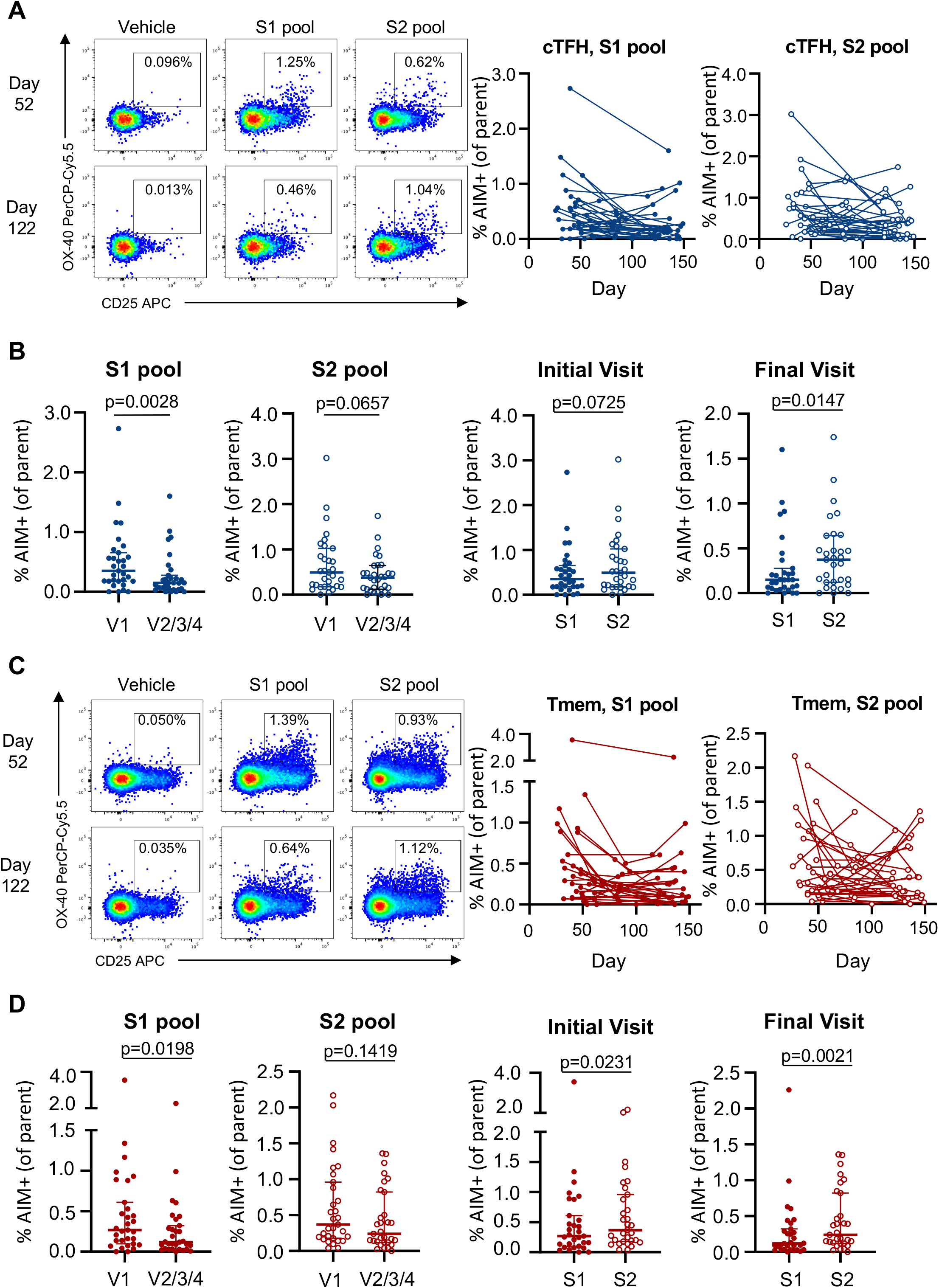
S1 and S2-specific CD4+ T cell responses. **(A, C)** Representative staining of AIM markers following S1 and S2 peptide pool stimulation among **(A)** cTFH (CD3+CD4+CD8-CD45RA-CXCR5+) or **(C)** CD4+ Tmem cells and longitudinal cohort analysis (n = 31). **(B, D)** Comparison of S1 or S2-specific **(B)** cTFH or **(D)** CD4+ Tmem responses at the earliest and latest visit for each participant, as well as paired frequency of S1 versus S2 responses at the initial or final visit (n = 31). Statistics assessed by two-tailed Wilcoxon test.

**Extended Data Figure 10:**
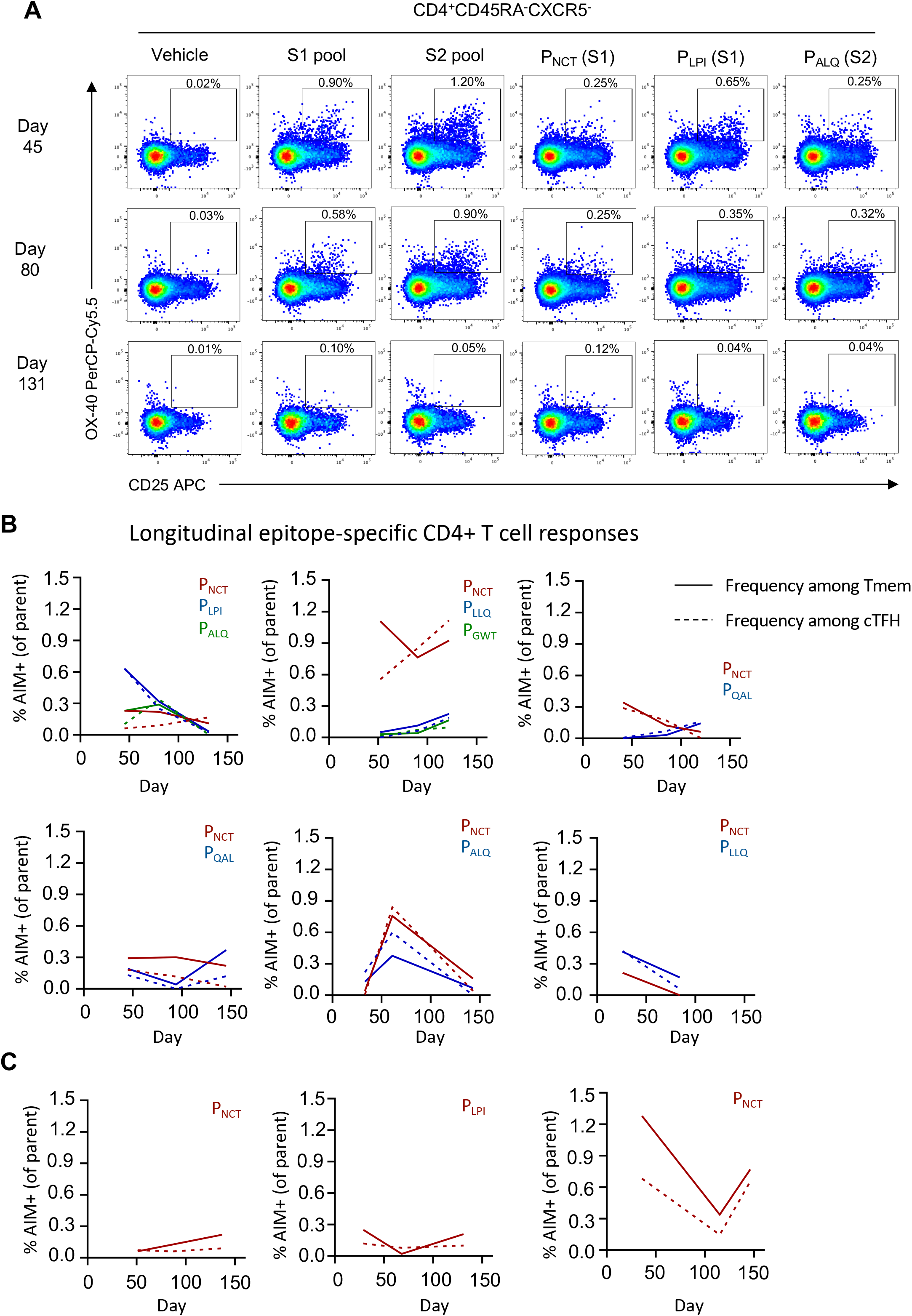
Epitope-specific CD4+ T cell responses. **(A)** Representative staining of AIM markers following S1 or S2 peptide pool or individual peptide stimulation among the CD4+ Tmem population. **(B,C)** Longitudinal peptide-specific frequencies in individual subjects (n = 9; solid line, CD4+ Tmem; dashed line, cTFH) for whom **(B)** multiple or **(C)** single epitopes were identified.

